# User adherence and perceptions of a Volunteer-Led Telemonitoring and Teleorientation Service for COVID-19 community management in Peru: The COVIDA project

**DOI:** 10.1101/2023.06.08.23291037

**Authors:** Stefan Escobar-Agreda, Javier Silva-Valencia, Percy Soto-Becerra, C. Mahony Reategui-Rivera, Kelly De la Cruz-Torralva, Max Chahuara-Rojas, Bruno Hernandez-Iriarte, Daniel Hector Espinoza-Herrera, Carlos A. Delgado, Silvana M. Matassini Eyzaguirre, Javier Vargas-Herrera, Leonardo Rojas-Mezarina

**Affiliations:** Unidad de Telesalud, Facultad de Medicina de San Fernando, Universidad Nacional Mayor de San Marcos, Lima, Perú; Universidad Continental, Huancayo, Perú; Universidad Peruana Cayetano Heredia, Lima, Perú; Instituto Nacional de Salud, Lima, Perú

**Keywords:** telemonitoring, volunteers, adherence, COVID-19, Peru

## Abstract

**Background:** During the pandemic in Peru, the COVIDA project proposed an innovative way to provide telemonitoring and teleorientation to COVID-19 pandemics led by health student volunteers. However, it has not been described how this interaction is perceived from the user’s perspective. The aim of this study is to describe the adherence and perceptions of users about COVIDA.

**Methods:** A mixed-method study was conducted to evaluate the adherence and perceptions of COVIDA users. This telehealth intervention implemented in Peru from August to December 2020 involved daily phone-calls by volunteer students to monitor registered users for 14 days or until a warning sign was identified. The volunteers also provided teleorientation to address the users’ needs and concerns. Quantitative analysis described the characteristics of users and assessed the factors related to adherence to the service. Qualitative analysis trough semi-structured interviews evaluated the user’s perceptions about the service.

**Results:** Of the 778 users enrolled in COVIDA, 397 (54.7%) were female and had a mean age of 41 years (SD: 15.3). During the monitoring, 380 users (44.4%) developed symptoms, and 471 (55.5%) showed warning signs for COVID-19. The overall median of adherence was 93% (p25:36%, p75:100%). Among those users who did not develop warning symptoms, a high level of adherence (>66%) was seen predominantly in users that developed symptoms and those with a positive COVID-19 test (p<0.05). Users referred that the information provided by volunteers was clear and valuable and, their accompaniment provided them with emotional support. Communications via phone calls were developed fluently without interruptions.

**Conclusions:** COVIDA represented an affordable, well-accepted, and perceived alternative model for telemonitoring, teleorientation and emotional support from student volunteers to users with diseases such as COVID-19 in a context of overwhelmed demand for healthcare services.

## 1. INTRODUCTION

The COVID-19 pandemic has posed a significant public health challenge due to its widespread impact and capacity to develop severe cases and fatalities in the population (1). While timely medical care has been shown to prevent such outcomes (2), patients encounter challenges in seeking care due to difficulties in self-monitoring due to a lack of knowledge of their symptoms and the possibility of developing severe conditions even in the absence of symptoms (3).

In this context, telemonitoring has emerged as an essential alternative to facilitate the follow-up of COVID-19 patients using Information and Communication Technologies (ICT) (4). In Peru, one of the countries most affected by this disease worldwide (5), the government implemented some institutional initiatives to promote remote monitoring to these users (6). However, despite their high demand or services, the implementation of these interventions was limited by the lack of availability of health workers, including their losses during pandemics (7).

Considering the willingness of health sciences students to participate in health interventions utilizing ICT (8) and provide assistance in the context of emergency (9), the Telehealth Unit of the National University of San Marcos (UNMSM) and the Peruvian National Institute of Health (INS-Peru), collaborated to design and implement the COVIDA Project. This intervention was a telemonitoring and teleorientation service carried out by university student volunteers and supervised by a team of healthcare professionals (10) to the prompt identification of severe signs related with COVID-19.

Volunteer-based telehealth services during the COVID-19 pandemic have been found helpful in other international experiences (11,12). However evidence in resource-limited settings is scarce, especially in the evaluation of the user perspective that would be crucial, to the development of patient-centred telehealth solutions, leading to improved adherence, better health outcomes, and increased satisfaction with healthcare services. This study aims to describe the adherence and users’ perceptions about their participation in the COVIDA Project in Perú.

## 2. METHODS

### 2.1. Study design

A mixed method study, with an explanatory sequential design was conducted, as is considered best practice in evaluating telehealth interventions (13). First, quantitative analysis was performed to gather information on user characteristics and their level of adherence to the service. Then, qualitative analysis was conducted to amplify the user and volunteer experiences providing deeper insights from both perspectives as users and providers.

### 2.2. The COVIDA Project

#### Design and services

The COVIDA project consisted of a telemonitoring and teleorientation service provided in Peru between September to December 2020 after the end of the first wave of infections of COVID-19 in this country. The telemonitoring component involved daily phone calls to identify those with the suspicion of developing severe COVID-19 symptoms based on warning signs such as shortness of breath, chest pain, persistent fever, cyanosis, or low oxygen saturation levels (less than 93% when the patient had a pulse oximeter) following the World Health Organization guidelines (14).

Sociodemographic and clinical information from participants was registered in virtual forms using a free software application called Kobo Collect ©. Additionally, volunteers provided teleorientation to educate users on adopting preventive behaviors, avoiding unsupervised medication, resolving their doubts and misconceptions about COVID-19, and guiding them to seek timely medical care when present warning signs. These calls were performed during the day according to the user’s availability and were expected to continue until complete 14 days after the start of their illness, which was determined by their symptom onset date or a recent positive test result, or until they develop a warning sign (See Figure 1).

**Figure 1.**
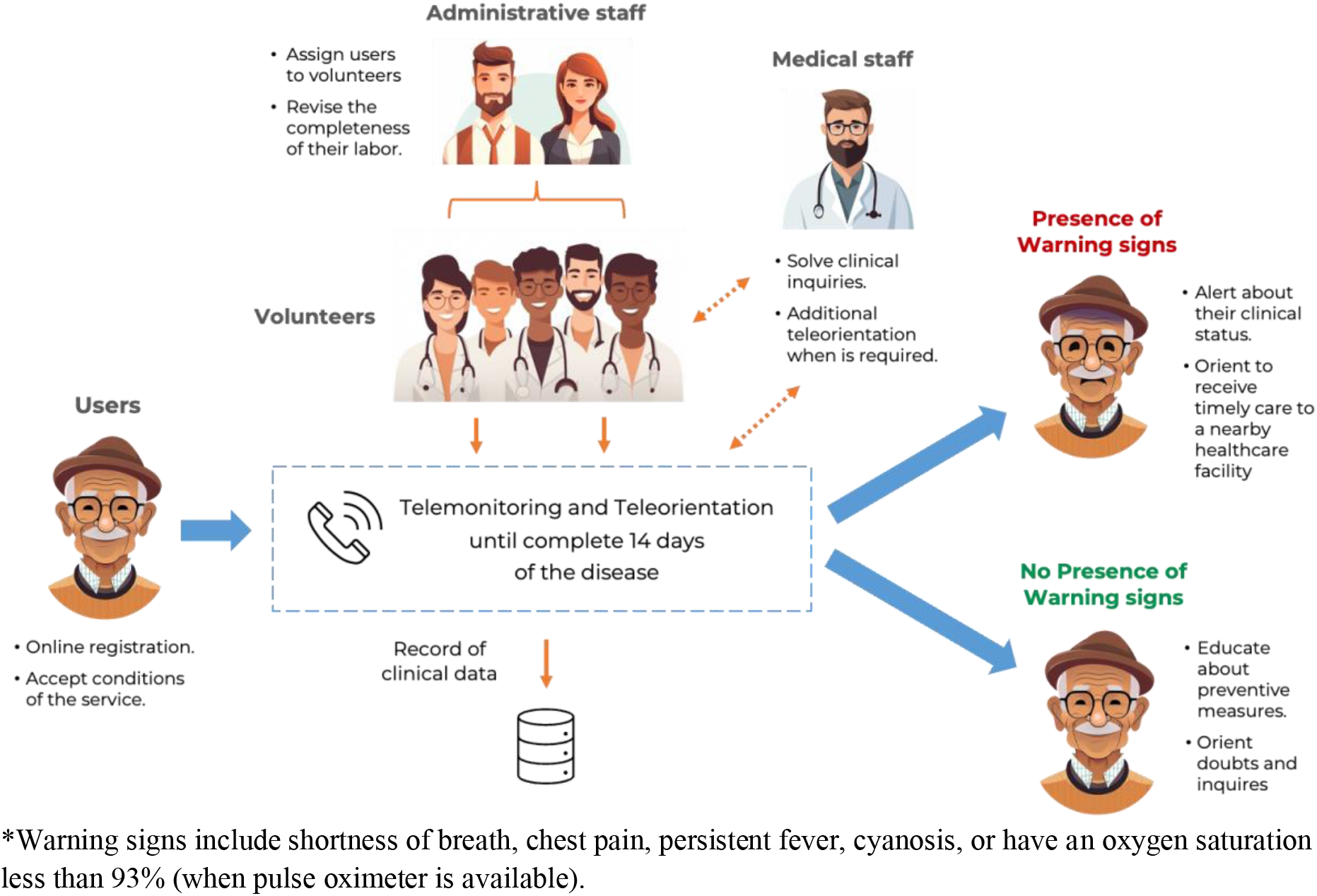
Design of COVIDA for the telemonitoring and teleorientation of COVID-19 in Peru.

#### Users, volunteers, and support team

The COVIDA Project was designed for adult users (≥ 18 years) in Peru who had tested positive for COVID-19, had related symptoms of the disease (see Table S1 Supplementary appendix), or had close contact with a positive case. The volunteers that provided the service were undergraduate or recently graduated health students who received training from INS-Peru on essential aspects of COVID-19, communication skills in emergencies, and informatics applications for calls and virtual surveys. Users and volunteers were recruited through social media, online newspapers, and the website of UNMSM and INS-Peru (10,15).

Additionally, this intervention counted a support team that included an administrative staff in charge of recruiting information of registered users, assigning to volunteers according to their availability and checking the realization of their labor, and a medical staff that oversaw the solving of inquiries made by volunteers and provide additional teleorientation to users in the case of complex cases or if the user would ask for it (See Figure 1).

### 2.3. Variables and data collection

For this study, we collected quantitative data from the telemonitoring registers of the COVIDA Project provided by UNMSM. We use these data to describe the characteristics of the users who participated in this service and their level of adherence. The characteristics evaluated included age, sex, region of origin, health insurance, presence of comorbidities, number of days of telemonitoring, development of symptoms, and development of warning signs. We defined the level of adherence as the percentage of days on which the user was effectively telemonitored according to what was planned for each case (See design of the COVIDA Project).

We also conducted semi-structured interviews with users to explore their experiences and perceptions about their participation in the COVIDA Project. The interviews were conducted after concluded the service via phone calls and lasted approximately 10 to 15 minutes. We focus on evaluating three main categories related to their experience: communicative, interaction, and technological aspects.

### 2.4. Analysis

#### Quantitative component

We performed descriptive analyses to describe the sociodemographic and clinical characteristics of the users who participated in the COVIDA project. Numerical data were summarized using means and standard deviation. For categorical data we used frequencies and proportions. Bivarate analyses were performed to compare the characteristics of users who developed warning signs with those who did not. We used Chi squares tests to compare categorical data and T Student tests to compare numerical data.

Multivariate analysis were performed to evaluate the characteristics of the users that would be related to a higher adherence to the service among those that did not developed warning signs. Considering that adherence is an outcome being bounded between 0 and 1, a fractional regression model was applied calculating Odds Ratio (OR) with 95% confidence intervals using crude, fully adjusted and parsimonius models. Detailed information about these analsys was described in Supplementary Appendix. All analyses were executed using Stata IC version 17.

#### Qualitative component

For qualitative analysis, we used a phenomenological approach. Five users who agreed to the interview were selected. Semi-structured interviews were conducted, recorded, transcribed, and compiled into a matrix for further analysis. Through the examination of the collected information, we sought to identify key themes and patterns that could provide a valuable understanding of the phenomenon studied.

### 2.5. Ethical aspects

This study was approved by the Social Security (EsSalud) Institutional Review Board for COVID (Ethics Certificate N° 96 – June 2020). Interviewed participants were enrolled after the completion of the informed consent application, and their information was managed confidentially only by the researchers of the study. The present study was carried out respecting the Helsinki and Taipei declaration research principles.

## 3. RESULTS

### 3.1. Quantitative results

From the 1794 people initially registered online to participate in COVIDA, 670 (37·3%) showed to have a telemonitoring history. In addition, 108 patients not initially registered also have shown to have a telemonitoring history giving a total of 778 users of this intervention. Users have a mean age of 41 years (SD:15·3) and were monitored for a median of 2 days (p25:1, p75:4). 424 (55·4%) were female, 475 (63·7%) were from Lima-Callao (the capital city), 528 (78·8%) reported having some form of health insurance, and 173 (22·2%) reported having at least one comorbidity. (Detailed clinical information is shown in Table S1 and S2, Supplementary appendix). Comparison of characteristics between those that developed and did not develop warning signs during telemonitoring was similar except for the presence of comorbidities that were more common in those that developed warning signs (p<0·05) (See Table 1).

**Table 1.** Characteristics of participants on the COVIDA project, overall and according to the development of warning signs during the telemonitoring. Peru, 2020.

|  | All users |  | With warning signs |  | Without warning signs |  | *p-value\** |
| --- | --- | --- | --- | --- | --- | --- | --- |
|  | Nro. | (%) | Nro. | (%) | Nro. | (%) |  |
| **Total** | 778 | (100.0) | 241 | (31.0) | 537 | (69.0) |  |
| **Age, mean (SD)** | 41.0 | (15.3) | 42.0 | (14.2) | 40.6 | (15.8) | 0.24 |
| **Sex** |  |  |  |  |  |  |  |
| Male | 341 | (44.6) | 98 | (41.2) | 243 | (46.1) | 0.2 |
| Female | 424 | (55.4) | 140 | (58.8) | 284 | (53.9) |  |
| **Macro-region of origin** |  |  |  |  |  |  |  |
| Lima-Callao | 475 | (63.7) | 137 | (59.6) | 338 | (65.5) |  |
| Center | 67 | (9.0) | 23 | (10.0) | 44 | (8.5) |  |
| North | 132 | (17.7) | 48 | (20.9) | 84 | (16.3) | 0.52 |
| South | 61 | (8.2) | 19 | (8.3) | 42 | (8.1) |  |
| West | 11 | (1.5) | 3 | (1.3) | 8 | (1.6) |  |
| **Health insurance** |  |  |  |  |  |  |  |
| Not | 142 | (21.2) | 44 | (21.1) | 98 | (21.3) |  |
| Yes | 528 | (78.8) | 165 | (78.9) | 363 | (78.7) | 0.95 |
| **Presence of comorbidities** |  |  |  |  |  |  |  |
| Not | 605 | (77.8) | 164 | (68.0) | 441 | (82.1) |  |
| Yes | 173 | (22.2) | 77 | (32.0) | 96 | (17.9) | <0.001 |

SD: Standard deviation, IQR: interquartile range
\*Chi-squared test and T-Student test.

Users in the COVIDA Project have a median adherence of 93% (p25:36%, p75:100%). Among those who did not develop warning signs, the factors related with a higher adherence to the service was, had comorbidities (OR: 1·81; 95%, CI: 1·28 - 2·56), had a recent positive test of COVID-19 at registration (OR:1·94, 95% CI: 1·48 - 2·55), or had COVID-19-related symptoms at registration (OR: 2·92, 95%CI: 2·16 - 3·95) (See Table 2).

**Table 2.**
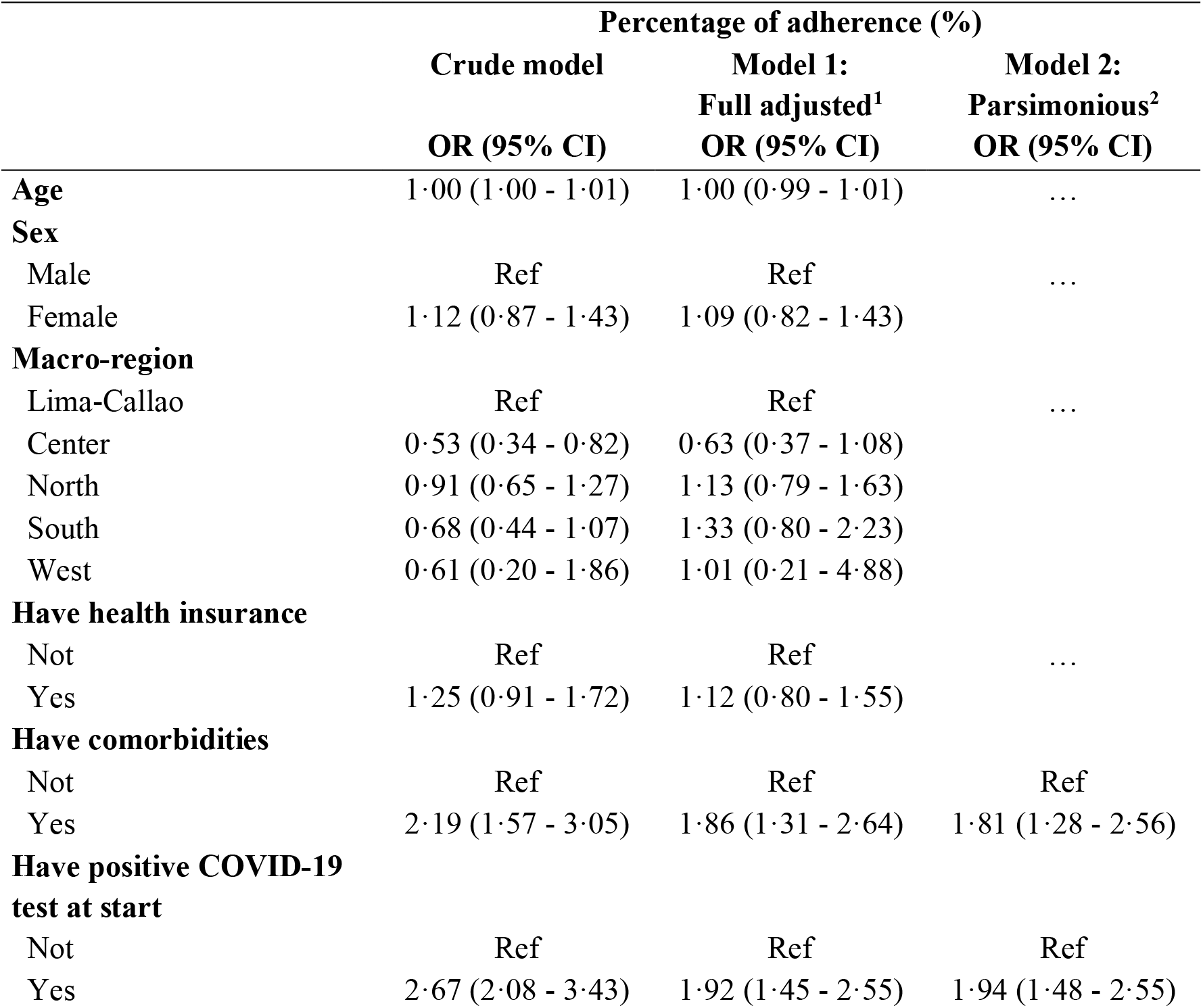
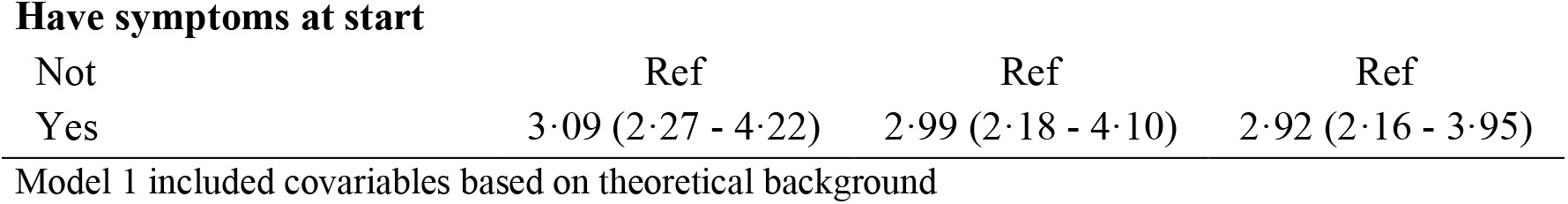
Relationship of percentage of adherence (%) and sociodemographic and clinical characteristics among those users who did not develop warning signs during the COVIDA project.

### 3.2. Qualitative results

On the communicational aspects, users reported that volunteers provided clear information using simple language. In addition, users highlighted their willingness, kindness, and patience to provide explanations. In some cases, volunteers became emotional companions, offering comfort during anxiety and sadness related to the illness.

> *“The volunteers attend you with patience and kindness, although I asked the same questions, they did not hesitate to explain you again and nice so that you understand (*….) *when I complained that I was worried they were also kind” (User 3)*
>
> *“The lady who attended me, apart from worrying about my physical health, listened to me, I think she has been an important support, they were difficult times, and that someone listens to you helps you a lot, so I am grateful” (User 2)*

The interactive aspects of the service posed some challenges, as some users were uncertain if the volunteer labor was supervised or if the volunteers were doctors. These initial misconceptions are supposed barriers to volunteer work.

> *“I knew they were medical students from San Marcos, but I didn’t understand how it worked, I think they are supervised by doctors, but it has been a good service” (User 1)*

In addition, some users report annoyance towards the frequent calls, including those made during nighttime hours, despite having initially agreed to this arrangement. This was more common in those asymptomatic or with milder symptoms.

> *“I know they wanted to help us, but sometimes they called you late and asked you the same thing again, that generated a bit of annoyance to me” (User 4)*
>
> *“I think that if you were not so serious, they should not call you so much, just every couple of days, because we are also busy or else, they should notify us in advance at what time they would call us” (User 5)*

Finally, regarding the technological aspects, users did not report any significant issues and stated they had smooth communication with volunteers without interruptions.

> *“I have been able to communicate fluently with the volunteers, there were no inconveniences in the phone calls*.*” (User 2)*.

## 4. DISCUSSION

The COVIDA Project is a volunteer-led telemonitoring and teleorientation service on COVID-19 that have shown great adherence and have been well-perceived by its users. One reason for this success is the fluent communication between users and volunteers facilitated by the simplicity of the technology used, which is based on phone calls. This approach has enabled the project to provide high-quality monitoring to users, regardless of whether they have access to pulse oximeters or not that have some considerations for its adequate use (16), have limited internet access commonly seen in countries like Peru (17) or do not have adequate digital literacy skills (18) for using other technologies as mobile applications, which have been used in other contexts (4). Indeed, none of the users interviewed in our study experienced any technical issues during the monitoring process.

Another factor that may have contributed to the high adherence to the COVIDA Project is the potential for teleorientation and accompaniment in addition to telemonitoring. Interviewed users mentioned that this service provided clear and useful information about COVID-19 and created a space for them to be heard and receive emotional support to encompass their hardship moments. These aspects could potentially contribute to a reduction in psychological distress experienced by COVID-19 patients, which is often linked to their uncertainty about the disease (19,20). This finding is supported by a study by Chakeri et al. in 2020, which showed that implementing an intervention focused on providing information and maintaining communication with COVID-19 patients can significantly reduce their level of stress and anxiety (21).

Low level of adherence in the COVIDA Project was most common among asymptomatic users and those without a confirmatory positive test for COVID-19. Qualitative data indicated that these users feel annoyed with the daily monitoring schedule, which increased the probability of dropout of the service. Furthermore, the presence of warning signs was most common among users with comorbidities. These findings suggest that future interventions should offer varying schedules of telemonitoring based on user preferences and should be focused on users with risk factors for the disease being monitored to increase adherence and effectiveness, respectively. Additionally, complementing remote monitoring with the use of medical devices, when available, could improve monitoring adherence in asymptomatic patients or those that would feel annoyed with frequent calls (22).

The participation of students as volunteers has a crucial role in the sustainability of the COVIDA project. Previous studies have shown that students are valuable resources for community health work due to their willingness to participate in such activities (11,23). During the first year of the COVID-19 pandemic in Peru, the availability of students was heightened as many educational activities in hospitals were suspended due to the high risk of SARS-CoV-2 infection (24). While other telemonitoring experiences have relied solely on the participation of health professionals (25), this experience demonstrated that these activities could, to some extent, be performed by students of health careers with proper training and supervision. This is especially important in settings with limited human resources for mass monitoring (7).

The participation of administrative and clinical staff was another important aspect of the sustainability of the COVIDA Project. As the telemonitoring and teleorientation activities were carried out voluntarily by students, the availability of some of them could be modified or suspended during the intervention affecting the continuity and quality of the monitoring provided. For this reason, the administrative staff was responsible for assigning patients to the volunteers, confirming their availability daily, and reassigning work to another volunteer if necessary. Additionally, the clinical staff clarified any doubts the volunteers had about using the virtual tools available in the intervention or aspects of COVID-19. They also provided additional teleorientation to users with complex cases of those requiring it. This support was essential to ensure the smooth running of the project and the provision of high-quality monitoring to users.

The evaluation of the COVIDA project has some limitations that need to be highlighted. First, there was a high rate of participants with no telemonitoring information from those initially registered. According to the data provided for analysis, some telemonitored users had no initial registration. This could be due to misregistration of the identification code of users. In COVIDA, user identification codes were the number of their National Identification Document (DNI) registered manually on each telemonitoring contact. Using virtual forms specialized in the follow-up of users could help prevent the loss of information in telemonitoring interventions caused by misregistration of identifiers. Despite these limitations, our study demonstrated that the COVIDA project was an affordable and well-accepted initiative in identifying and guiding people suspicious of developing a severe case of COVID-19 in Perú. The project was also a sustainable intervention in a context with an overwhelming demand of attention and limited economic and human resources.

The COVIDA Project has presented a promising model for providing telemonitoring and teleorientation services for COVID-19, which has been well-received and sustainable. Given its characteristics, it could be adapted to monitoring other diseases where a severe or fatal outcome should be prevented in the short term, such as diabetes or hypertension. However, further research is needed to evaluate the clinical effectiveness of such interventions in preventing severe outcomes like hospitalization, ICU admission, and death. Additionally, it is crucial to address scalability issues in future studies to ensure that similar interventions can be implemented on a larger scale and in other settings. Overall, the COVIDA Project has demonstrated the potential of telemonitoring and teleorientation in resource-constrained settings and highlights the need for continued research and innovation in this area.

## Supporting information

COVIDA Project

## Data Availability

The datasets analyzed during the current study are not publicly available as is being part a confidential information of patients be attended into the COVIDA Project but could be anonymized and shared on reasonable request.

## 6. ACKNOWLDEGMENTS

We would like to express our sincere gratitude to all the students and medical professionals who contributed as volunteers of the COVIDA project providing telemonitoring and teleorientation to participants. Your dedication, hard work, and vocation to help others have been instrumental in the success of this intervention.

