## Supplementary figures and images for "User adherence and perceptions of a Volunteer-Led Telemonitoring and Teleorientation Service for COVID-19 community management in Peru: The COVIDA project"

### COVIDA Project

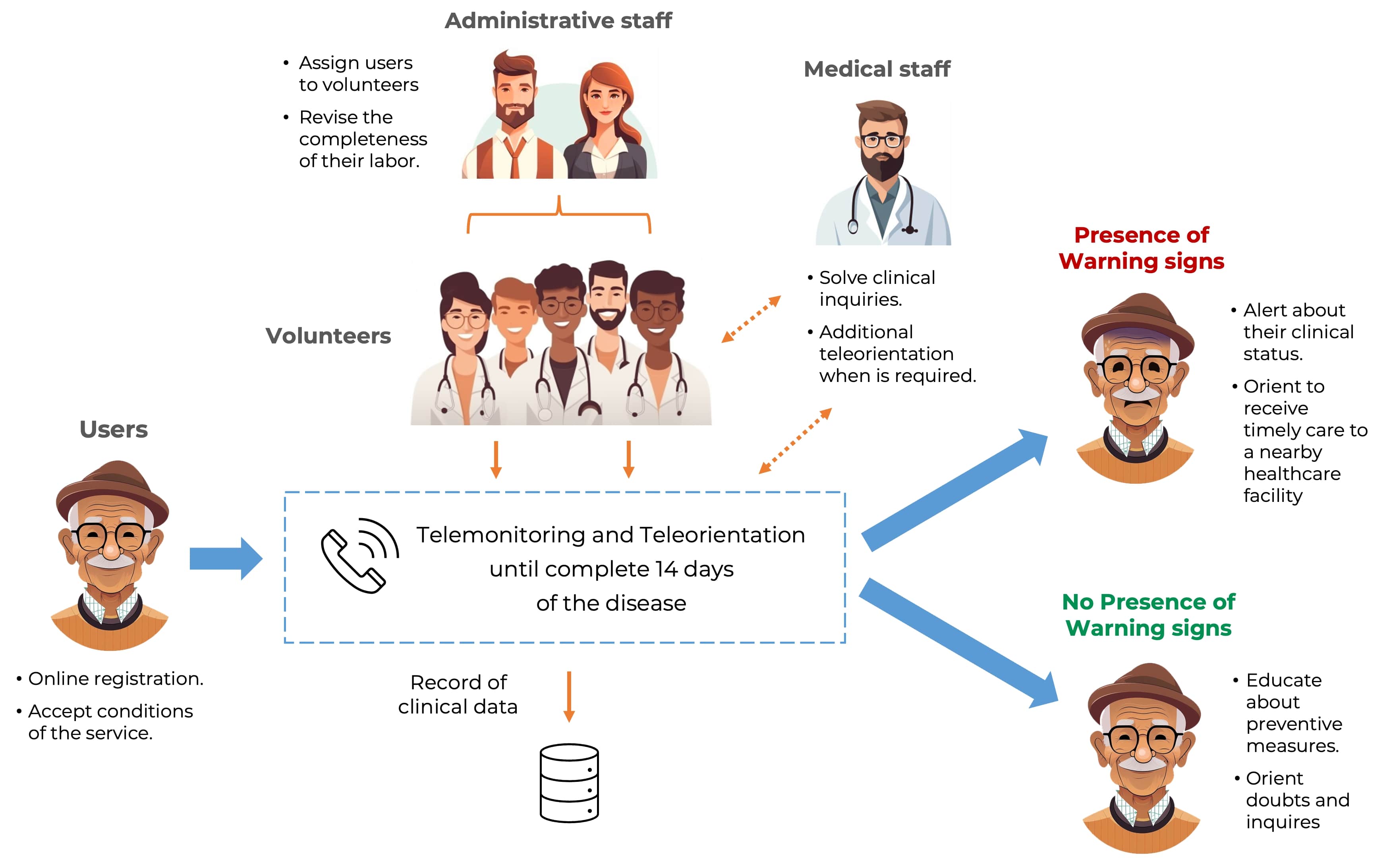
