## Supplementary Appendix for "User adherence and perceptions of a Volunteer-Led Telemonitoring and Teleorientation Service for COVID-19 community management in Peru: The COVIDA project"

### S1. Scope and diffusion of the COVIDA Project

In Peru, the COVIDA project was developed as an initiative in response to the increase infected, hospitalized, and deceased people by COVID-19, during 2020. Although the implementation of this project was carried out during a period of low incidence of cases after the culmination of the first wave of infections in Peru, came to have an important scope at the local level given the diffusion it received through various local media (1) as well as the institutional support of the Universidad Nacional Mayor de San Marcos (UNMSM) and the National Institute of Health of Peru (2,3).

In the same way, the important participation of volunteers in COVIDA, was facilitated by the call for social networks carried out by the UNMSM and local media, in addition to the network of volunteers previously established by the university, the development of "Tele-triage" another initiative organized at the beginning of the pandemic to the identification of suspected cases of COVID-19 that was deployed with the participation of student volunteers (4). Both the dissemination activities as well as the organization and training of volunteers were carried out virtually and remotely given the context of mandatory social isolation in the country due to the pandemic. Despite this, all these activities were carried out successfully.

### S2. Informatic resources for telemonitoring and teleorientation

To facilitate telemonitoring and teleorientation activities by COVIDA volunteers, some informatic resources were available. For telemonitoring, there was a computer application that allowed massive and anonymous calls to patients. In this way, the volunteers could contact the patients continuously, without the need to use their personal numbers, respecting their privacy and preventing patients from contacting them outside this intervention. Regarding the register of information collected in telemonitoring, the Kobo Collect application was used, which is free to use, allows a large number of registers to be stored and can be used off-line if there is limited or no nearby Internet connection (5).

Despite its benefits, the Kobo Collect application lacks an automated identification feature for participants undergoing follow-up, which requires volunteers to manually enter the national identity document (DNI) number of each participant in every record. As a solution, the CommCare application was utilized during the final weeks of the intervention to serve as the registration platform for COVIDA. By generating random codes as unique identifiers for each participant, CommCare minimizes the risk of errors that could occur when volunteers enter this information manually (6).

Another important virtual resource used during this intervention was the COVIDA website, a virtual space that included a collection of various documents, links, and sources of information of interest that the volunteer could use to provide guidance and/or share it with the monitored patients. This included, for example, basic concepts about COVID-19, current local regulations about COVID-19, and measures to prevent the spread of this disease in symptomatic patients.

### S3. Characteristics of participants

#### *Region and health insurance*

Regard to the region of origin of users in the COVIDA Project, most of them came from Lima (459, 59.0%), Lambayeque (122, 15.7%), and Puno (29, 3.7%).

**Table S1. Health Insurance of users of the COVIDA Project, Peru**

| | n | (%) |
|---|---|---|
| **Total** | 778 | (100.0) |
| **Region** | | |
| Amazonas | 2 | (0.26) |
| Ancash | 24 | (3.08) |
| Apurimac | 4 | (0.51) |
| Arequipa | 6 | (0.77) |
| Ayacucho | 11 | (1.41) |
| Cajamarca | 3 | (0.39) |
| Callao | 16 | (2.06) |
| Cusco | 17 | (2.19) |
| Huancavelica | 3 | (0.39) |
| Huanuco | 3 | (0.39) |
| Ica | 8 | (1.03) |
| Junin | 10 | (1.29) |
| La Libertad | 4 | (0.51) |
| Lambayeque | 122 | (15.68) |
| Lima | 459 | (59.0) |
| Loreto | 3 | (0.39) |
| Madre de Dios | 4 | (0.51) |
| Moquegua | 6 | (0.77) |
| Pasco | 4 | (0.51) |
| Piura | 3 | (0.39) |
| Puno | 29 | (3.73) |
| San Martin | 2 | (0.26) |
| Tacna | 3 | (0.39) |
| No information | 32 | (4.11) |

Among the 528 users that have reported to have health insurance, most common was Social Security (271, 40.45%), and Integral Health Insurance (SIS) (213, 31.79%)

**Table S2. Health Insurance of users of the COVIDA Project, Peru.**

| | n | (%) |
|---|---|---|
| **Total** | 528 | (100.0) |
| **Health Insurance*** | | |
| Social Security (EsSalud) | 271 | (40.45) |
| Integral Health Insurance (SIS) | 213 | (31.79) |
| Private | 35 | (5.22) |
| PNP | 16 | (2.39) |
| Armed Forces | 5 | (0.75) |

**Row percentages*

*Clinical characteristics*

As part of the monitoring carried out in COVIDA, various clinical characteristics of the participants were recorded, including the presence of symptoms associated with COVID-19 as well as the presence of warning signs. Among the most common symptoms manifested during monitoring was the presence of cough in 303 people (38.9%), hyposmia or anosmia in 292 people (37.5%), and headache in 276 people (35.5%).

**Table S3. Symptoms reported by users in the COVIDA Project, Peru.**

| | Nro. | (%) |
|---|---|---|
| **Total** | 778 | (100.0) |
| **Identified symptoms*** | | |
| Cough | 303 | (38.9) |
| Hyposmia/Anosmia | 292 | (37.5) |
| Headache | 276 | (35.5) |
| Sore throat | 245 | (31.5) |
| Muscular pain | 231 | (29.7) |
| Congestion | 176 | (22.6) |
| Fever | 137 | (17.6) |
| Diahrrea | 87 | (11.2) |
| Articular pain | 71 | (9.1) |
| Abdominal pain | 54 | (6.9) |

**Row percentages*

Regarding the warning signs identified, shortness of breath and chest pain were identified among the most common signs. The evaluation of oxygen saturation through pulse oximeters was carried out in those patients who previously had this device, and in some others to whom it was distributed as part of the project due to the presence of risk factors for COVID-19 such as the presence of comorbidities or in elderly patients who may be more prone to developing a severe condition of the disease.

**Table S4. Warning signs identified in users in the COVIDA Project, Peru.**

| | Nro. | (%) |
|---|---|---|
| **O2 saturation (%)*** | | |
| ≥93% | 358 | (94.2) |
| <93% | 22 | (5.8) |
| **Type of warning signs** | | |
| Shortness Of Breath | 90 | (57.0) |
| Chest Pain | 38 | (24.1) |
| Persistent Fever | 25 | (15.8) |
| Cyanosis | 5 | (3.2) |

### S4. Regression models for multivariate analyses

To evaluate the relation of user`s characteristics of the COVIDA Project and their adherence to this service we used a fractional regression model considering that adherence, as the outcome variable, resulting in values between 0 and 1 enabling to estimate of odds ratios of these relationships for each predictor evaluated.

The fractional regression model was parameterized through a generalized linear model (GLM) with a binomial family distribution, a logit link function, and robust standard errors, as estimated by the Huber-White sandwich estimator. Theoretical considerations guided the selection of predictor variables, generating a comprehensive model.

To identify a more parsimonious model and ascertain the most suitable form for the single continuous predictor, age, a backward variable selection process was implemented as part of the multivariable fractional polynomial modelling approach utilizing the “mfp” command in Stata. The level of significance for form selection was set at 0.1. Age, as a numerical variable in its original linear form, proved to best fit the data. Moreover, a normal quantile-quantile plot of the quantile residuals did not disclose any significant deviations from linearity within the comprehensive model. Wald-type confidence intervals were reported at a 95% level.

### S5. Bibliographic references
